# Music education reduces emotional symptoms in children: A quasi-experimental study in the Guri Program in Brazil

**DOI:** 10.1101/2024.11.12.24317159

**Authors:** Graziela Bortz, Beatriz Ilari, Nayana Di Giuseppe Germano, Andrea Parolin Jackowski, Hugo Cogo-Moreira, Patrícia Silva Lúcio

## Abstract

The transferability of music education to cognitive and social skills has been explored recently, but its causal effects remain debatable. This quasi-experimental study aimed to examine the impact of a music education program (Guri) on aspects attention, working memory and socioemotional skills in children from underserved communities of São Paulo, Brazil. The music group (n = 38, 5-9 years) was recruited from 10 centers (polos) distributed across the metropolitan area of São Paulo and the control group (n = 67) consisted of aged-matched children who attended public schools surrounding the polos. The ABEP SES questionnaire and the Raven test were used as control variables, the SDQ questionnaire, the Digit-span subtest from WISC-IV, the PARB-Q questionnaire, and the BPA test were used as subject variables. A significant effect of the intervention in emotional symptoms in the music group as opposed to controls was found (F(1,89) = 4.562, p = 0.035, η^2^ = 0.049), indicating benefits for the music group. Children whose mothers had low levels of education benefited with gains between 0.70 and 0.95 standard deviations in both groups for divided attention, indicating a significant interaction with maternal education.

## Introduction

### Music education and executive functions

The hypothesis of the effects of music education practice on cognitive abilities and brain plasticity, with potential far transferability to other domains, has been the focus of several studies published in the last decade (1–11). While some studies suggest that abilities related to executive functions (EFs) are particularly enhanced by musical training (2–4), others remain inconclusive (11, 12). Yet inhibitory control, one of the central dimensions of EFs, appears to be a consensus in terms of its transferability from music education/music training, especially among preschoolers (12).

EFs consist of a set of abilities that promote goal-directed and autonomous behavior, and are crucial to stablish day-to-day goals as well as more complex and long-time planning (13–17). Although validated models of EFs are still controversial (14, 18, 19), there are commonalities among proposed theoretical models, including dimensions such as attention, inhibitory control, working memory (WM), fluid intelligence, problem solving, decision making, and cognitive flexibility.

EFs can be classified as *hot*, or those associated with emotional and social cognitive aspects relevant to decision-making process, or *cold*, which are linked to mental abilities involved in inhibitory control, WM, short-term memory (STM), reasoning and planning (20, 21). Cold EF tasks require mental effort and constant top-down supervision of the pre-frontal cortex (PFC) to prevent automatic actions (16).

According to Diamond’s model (17), and despite belonging to EF cold domains, attention, a subdomain of inhibitory control, can be affected by emotional and cognitive self-regulation (22, 23). Attention requires effort and volition and, therefore, can also be involved in both hot and cold EFs depending on the type of stimulus to be inhibited (*e.g.*, self-control in delayed gratification or resistance to a distractor in attentional focus). The two types of attention (*i.e.*, emotional and cognitive) may not necessarily fall under the same factor, but are strongly correlated (17, 24). Petersen and Posner (23) updated their model of attention (22) to include three major networks: alerting, orienting, and executive networks. The latter further includes focal attention and regulation of processing networks.

Attention can also be classified according to the type of task at hand: (1) sustained attention/focused (*i.e.*, maintaining focus on a single stimulus/task over time), (2) divided attention (*i.e.*, concentrating on two or more stimuli/tasks simultaneously for a given time), and (3) shifting/alternated attention (*i.e.*, change the focus of the attention to the perceptual attributes of the stimuli under relevant stimulus-response maps rules) (25, 26). Attention also plays an important role in WM, another dimension of cold EF that requires processing and storage, sharing cognitive resources from both (27). Therefore, it is necessary to maintain a focus of attention by inhibiting the preponderant stimulus to prioritize information and allow updates of the environment, and subsequently coordinating them with automated responses in the use of WM. All this happens while new associations are created in STM.

A meta-analysis by Talamini et al. (8) found a large musician advantage for STM and WM with tonal stimuli, a moderate effect with verbal stimuli, and a non-significant effect with visuospatial stimuli. The systematic review of Rodriguez-Gomez and Talero-Gutierrez (12) examining the associations between WM and music training revealed three studies among eight that showed improvement in one subdomain (*i.e.*, either visual or spatial) following a music intervention with children. Additionally, in four studies reviewed by the authors, music education was found to improve selective attention among children, although only one study showed statistical significance. Furthermore, out of nine studies conducted with preschool-age children, six presented improvements in inhibitory control following music education. From these studies, one can assume that verbal WM may be a strong candidate for music education effects, as seen with selective attention.

### Hot executive functions, social competence, and social skills

Hot EFs are far less researched than cold EFs, even though psychometric instruments that consider level of affection, emotion self-regulation, affect decision making, empathy, and delayed gratification are often used in studies (15). One of the reasons for the lower incidence of articles on hot EFs might be the entanglements between the concept of hot EFs and the constructs of social competence, social skills and self-regulation. For Zimmerman et al. (20), “hot executive functions entail cognitive abilities supported by emotional awarness and social perception (*e.g.* social cognition).” The authors adapted their concept from the models of Chan et al. (28), and McDonald (29), placing social cognition on one side, and emotion cognition and decision making on the other, and subsequently positioning all under the umbrella of hot EFs. Social cognition, in turn, was also further divided into cold (*i.e.*, cognitive empathy and theory of mind) and hot (*i.e.* affective empathy and theory of mind, and emotion recognition) aspects (20). For example, while one person may make decisions based on *understanding* other people’s cues, others may arrive at decisions based on their *affections*. Unsurprisingly, both cold and hot social cognition have neural correlates (30–32).

Social competence has been referred to as the ability to maintain positive relationships while persuing personal goals (33). For Hukkelberg et al. (34), whereas social competence is a broad term, social skills can be situated within more specific behaviors, as the latter can be examined through observable variables that compound a social competence factor (35–38).

Both social skills and social competence are positively correlated with academic achievement and problem solving skills (35, 38, 39), and have been associated with positive mental health (35, 38). Social skills can be measured with scales and subscales that address cooperation, self-control, internalizing and externalizing problem behaviors, empathy and hyperactivity (35, 38, 39).

### Social skills and music

Associations between emotion self-regulation, prosocial skills, and music are still under scrutiny. While some studies observed effects of musical interventions on prosocial skills in preschoolers and school-aged children (40–42), others found moderate to null effects (11, 43, 44). Still other studies found effects of music education only on selected social skills of children and youth. Villanueva et al. (45), for example, found effects of formal music instruction on children’s sharing behaviors, theory of mind, and state empathy. Alemán et al. (11), in turn, found a significant improvement in behavioral difficulties detected via parental questionnaires and scales, especially among male children who had experienced violence and had mothers with low levels of formal education. Also using a parental scale, Ilari et al. (46) found a significant effect of four years of music education on children’s aggressiveness and hyperactivity. Similarly, Boal-Palheiros and Ilari (38) found effects of one year of music education on externalizing behavioral problems in Portuguese children from underserved communities. Thus, current evidence is mixed and needs further substantiation, and this is particularly true in the case of children from non-WEIRD (i.e., non-White, from English speaking, industrialized, rich and democratic) populations (47) and those at risk for behavioral problems.

### The present study

This study aims to contribute towards the area of music education through an examination of its impact on specific cognitive and socioemotional outcomes among children at risk for behavioral problems. This quasi-experimental investigation adds to the body of knowledge derived from cross-sectional and correlational research that found evidence supporting beneficial effects of music education in the target outcomes. Particularly, we focus on aspects of attention (shifting, divided, and sustained) and aggressive behavior, by comparing test results from a group of children who participated in formal music classes to matched controls.

As noted, most of the studies examining the associations between music, EFs, and social skills were conducted in the global North, predominantly in North America and Western Europe and with WEIRD populations (47). This study^1^ aimed to investigate whether participation in a musical program called *Projeto Guri* (48) designed to serve low-income communities in the city of São Paulo, Brazil, conferred any benefits to children’s cognitive and social skills, including those who were at risk for behavioral problems.

## Method

### Ethics

All research protocols were approved by the local Ethical Committee *Plataforma Brasil* n. 15049919.8.0000.5420, and the Rebec n. U1111-1243-8071. Written consent was also obtained from the municipality of São Paulo. A Memorandum of Understanding was created between the State University of São Paulo (Unesp) and the Guri Program, outlining the specifics of the research, expectations, and obligations from all involved parts.

### Sample and recruitment

Children and their families were invited to participate in the study through their music programs (Guri *polos*) and local schools. Due to ethical reasons related to the policies of enrollment in the Guri Program, it was not possible to randomize child participants into music and control groups.^2^ Therefore, we performed an intervention study based on a quasi-experimental design, which took place in 2022 during the Brazilian academic year—February through December.^3^ The researchers responsible for the statistical analyzes were blinded to the participants. The examiners were also blinded to those who analyzed the outcomes. The target music group consisted of children aged 5 to 9 years,^4^ who were about to start music classes for the first time in the Guri Program (48). This Program consists of a network of various centers –called *polos*– distributed across the metropolitan area of São Paulo, also known as *Grande São Paulo*. Most of the *polos* are located in the outskirts of the city and function within a public system of integrated schools called *Centros Educacionais Unificados* (Unified Educational Centers) or CEUs. Children in the music group were recruited from 10 different *polos*: 7 from the extreme east part of the city, which is also the most populated, 3 from the extreme south, and 2 from the extreme north. Beginning music classes take place weekly for one hour through a modality known as *musicalização* (49), a form of general music that is carried out through collective singing, movement, and performance of rhythms and melodies, using small percussion instruments, recorders, and Orff xylophones and metallophones.

The control group consisted of aged-matched children who attended regular schools surrounding the Guri *polos*. Exclusion criteria for sample recruitment in both groups were: 1) children had attended regular school music classes or instrumental/voice lessons before, and 2) children had received a diagnosis of ASD (Autism Spectrum Disorders) or ADHD (Attention Deficit Hyperactivity Disorder). Children in the experimental group were recruited at the time of enrollment in the Guri Program, when parents/guardians were informed about the study and given consent forms. After signing consent forms, parents were invited to complete the SDQ–parent version (50, 51) and the ABEP questionnaire (52) that assesses socioeconomic status. This procedure constituted the first screening phase and aimed to identify children who would attend the Guri Program in 2022, and who were at risk for behavioral problems in four of the five areas evaluated by the SDQ, namely, emotional symptoms, conduct problems, hyperactivity, and relationship problems.

Recruitment was done upon securing authorization from the school boards to participate in the research. The researchers met with parents at the schools to explain the study, get consent, and subsequently distribute questionnaires. When meeting at schools was not possible, questionnaires were sent home by children and subsequently collected. A total of 255 parent protocols were collected at the Guri Program and in schools surrounding the programs for the control group, out of which 137 protocols were excluded. After further screening with the Brazilian version of the Raven’s test (53, 54), children at risk for intellectual difficulties were excluded from the sample to avoid confounders (n = 10). One child was excluded for having previous music lessons and another was diagnosed with autism, totaling 149 exclusions. The total of participants (n = 106) consisted in the intervention group –music group (n = 45)– and the control group (n = 61). The music group lost 7 children, who dropped out of the music program at the follow up (FUP). Therefore, 38 children remained in the music group. One child was excluded of the control group at the FUP for late autism diagnosis.

Baseline data were collected in March and April of 2022, and the FUP tests and questionnaires were taken seven months later^5^. Because children differed significantly in both groups (see Sample Matching below), 35 children from the music group and 57 children from the control group were included in the final statistical analysis after matching (Fig 1).

**Fig 1.**
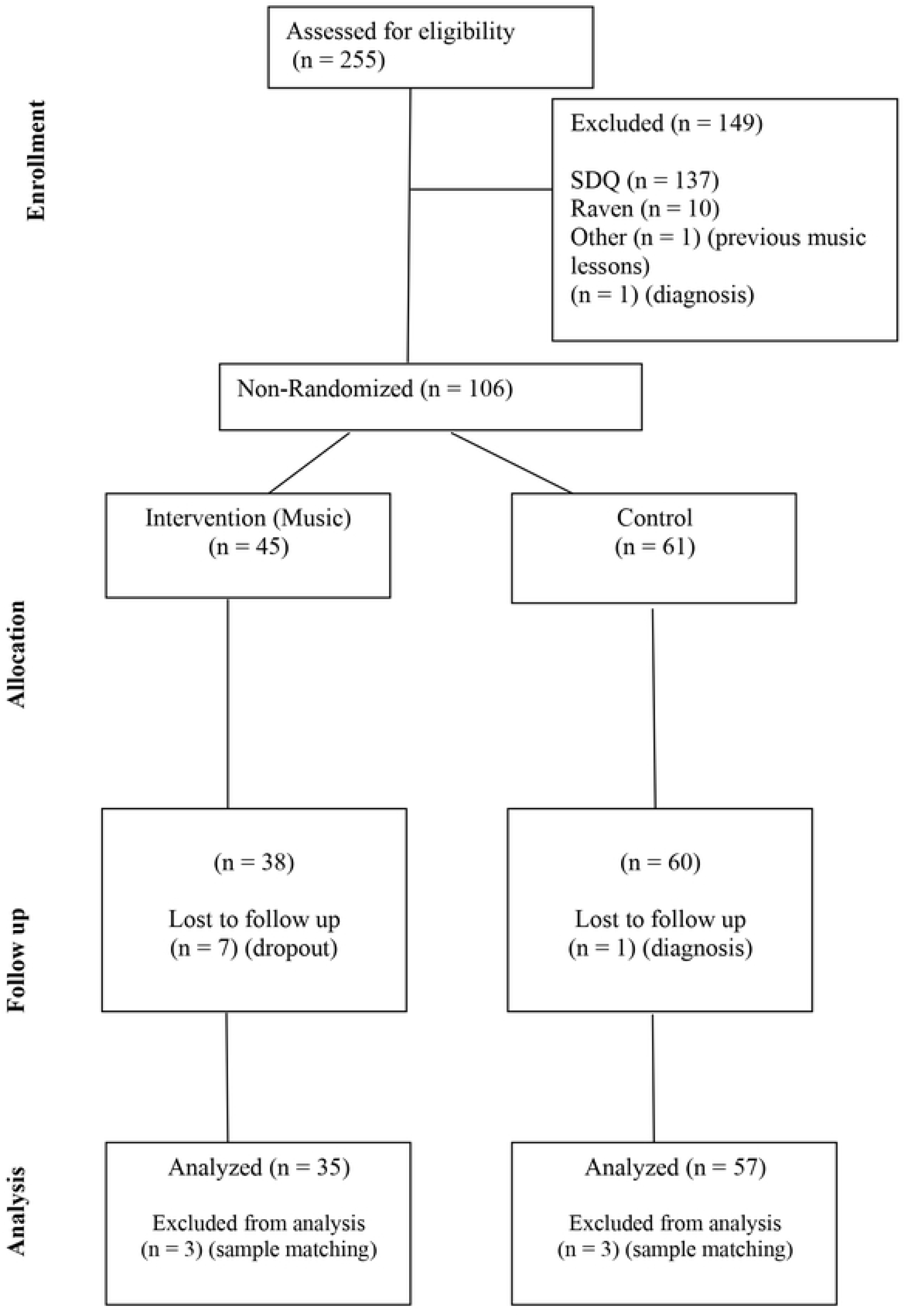
Flowchart of participants through each stage of the trial.

Children in the music group were 5-9 years old (mean = 6,61; S.D. = 0,72) and 52,6% boys at the baseline. In the control group, children were mostly boys (70%) and were 5-7 years old (mean = 6,28; S.D. = 0,49). Seven children from the music group dropped out from the research. Children who dropped out did not differ from those who completed the FUP in terms of sex (*χ*^2^(1) = 0,007, *p* = 1,000) or group assignment (*χ^2^*(1) = 2,321, *p* = 0,148), but they tended to be older at the baseline (*t*(96) = 2,589, *p* = 0,011). Two children were later diagnosed, one from the music group and another from the control group.

### Instruments

#### Control variables

- *Maternal schooling*: We used the Brazilian questionnaire of economic classification criteria –*Associação Brasileira de Empresas de Pesquisa* (52), created to assess socioeconomic status based on samples of households. This questionnaire contains nine items that assess: a) possession of durable consumer goods; b) type of water supply system and street paving; c) number of residents in the household; d) family composition; and e) level of education of the head of the family. The result of the questionnaire is a measure stratified into five socioeconomic classes, namely: A (subdivided into A1 and A2), B (also subdivided into B1 and B2), C, D and E. For the purpose of this study, we used the mother’s years of schooling measured by this questionnaire.
- *The Raven’s Colored Progressive Matrices* (CPM)–Special Scale (53, 54). We used this test to assess non-verbal fluid intelligence, analogical reasoning, and the ability to generate responses from non-verbal stimuli in participating children. This test is composed by 36 items dichotomously corrected (score of 0 for incorrect and 1 for correct answers), with a total score of 36. Normative data is given in percentiles, attributed by age group. Children with non-verbal intelligence falling under percentile 25 were excluded from the sample.

#### Subject variables

- *Strengths and Difficulties Questionnaire* (SDQ)–parent version (50, 51), Brazilian version (55). The SQD provides an assessment of behavior, emotions, and relationships of children and adolescents. Questions are grouped into five dimensions: emotional symptoms, conduct problems, hyperactivity/inattention, peer relationship problems, and prosocial behavior. Each item is scored between 0 to 2, according to respondents’ agreement with the statements (*e.g.*, if the mother/parent thinks that the statement “Restless, overactive, cannot stay still for long” is certainly true about his/her kid, the item is scored with 2 for the hyperactivity-inattention dimension). Some items present opposite directional relationship with the construct and must be reversed before scoring. Each dimension is scored on a scale of 0-10. The higher the score, the higher the behavioral difficulty or prosocial behavior. Difficulty items are summed up into a difficulty score varying from 0 to 40. For selecting children at behavioral risk, we used the cutoffs of difficulties scores ≥ 10, but the prosocial behavior scores varied across the sample.
- *Digit-span subtest for children from the WISC-IV* (56, 57). In this test, children are presented with increasingly long strings of numbers, composed by 2 to 9 items each (*e.g.*, 2-9-7). Upon hearing the sequence, the child must repeat the digits in the same order given (short term memory task) or in inverse order (working memory task). Each sequence size presents two different trials, and the child receives a score of 1 for each sequence correctly uttered. The total score in each task is 16. The test is discontinued when the child makes an error on two trials in a same string of numbers. In this study, we used the span score in reverse order, or the longest sequence numbers kept by the child in memory, as this subtest has been recognized as a robust measure of working memory (58).
- *Peer Aggressive and Reactive Behaviors Questionnaire* (PARB-Q/QCARP) (59), Brazilian version (60). This self-report questionnaire measures behaviors in terms of active and reactive aggressions among pairs (59). This instrument contains 20 questions and contains two scales: the Peer Aggression Scale (PA) and the Reaction to Peer Aggression Scale (RPA). The PA consists of five items that assess proactive (or instrumental) physical and verbal aggression (*e.g*., kicking or slapping) and three social-desirability bias items that are not scored (*e.g*., telling jokes). The questions begin with “how often do you…,” followed by the circumstances to be answered. The RPA consists of 12 sentences containing different ways reported by children about how they react to aggression from their peers. Questions like “when a colleague of yours…,” are followed by statements, which may signal reactive aggression (RA; 6 items, *e.g*., “hit or push you,” “do you hit your colleague?”), seeking teacher support (STS; 3 items, *e.g*., “hit or push you, do you tell the teacher?”), as well as internalized emotional reactions (IR; 3 items, *e.g*., “hit or push you, do you cry and sulk?”). Each question is rated on a 4-point Likert scale (every day, sometimes, a few times, and never). PA scores range from 0 to 15, and RPA scores range from 0-18 in the RA scale and 0-9 in the STS and IR scales.
- *Bateria Psicológica para Avaliação da Atenção* (BPA; Psychological Battery for Attention Assessment) (61). The BPA is a pencil-and-paper test divided into three subtests that assess different domains of attention: sustained attention (in one object among distractors), divided attention (focus in three objects among distractors), and alternated attention (alternating focus between different objects among distractors).

The entire test battery was completed in 40-50 minutes, depending on each child’s performance. Children were allowed to take breaks during the testing sessions.

### Sample matching

For the baseline measures, the groups differed significantly in terms of age, reactive aggression (QCARP), non-verbal intelligence, and mother’s years of schooling. As depicted in S1 Table, children in the music group tended to be older, presented higher levels of reactive aggression and non-verbal intelligence, and their mothers had higher levels of education. Moreover, even though sex did not significantly differ between the groups, the proportion of boys was higher in the control group. As the groups differed in terms of these characteristics, comparisons of the effects of music on children’s behavior would be biased. Therefore, we managed the sample characteristics to match study groups.

Sample matching was carried out based on the removal of children from both groups considering simultaneously the following criteria, respectively, in increasing order of importance: (1) older children from the experimental and younger from the control group; (2) higher levels of reaction to aggression in the experimental group and lower in the control group; (3) superior and average superior performance in the Raven test in the experimental group and lower performance in the Raven test in the control group. Given that mother’s years of education is not a subject variable but rather related to it, we opted for removing children whose mothers had more years of schooling from the experimental group, and the opposite in the control group. Following this procedure, we removed 6 children (3 from each group) and their results in these variables are depicted in S2 Table. With the matching, the final sample was composed by 92 children (experimental = 35; control = 57). Table 1 shows the comparison between groups in the baseline measures after matching.

**Table 1.**
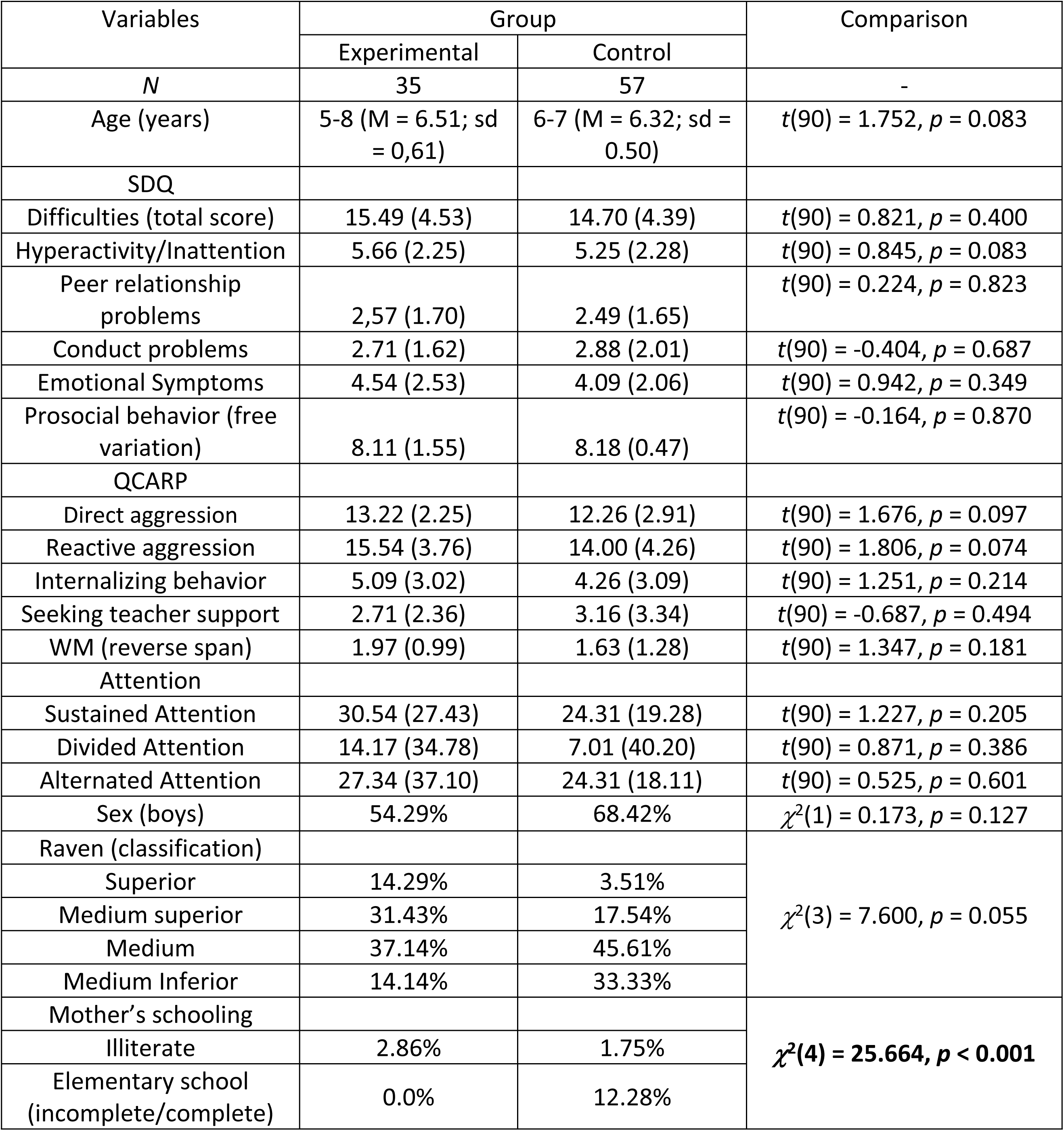
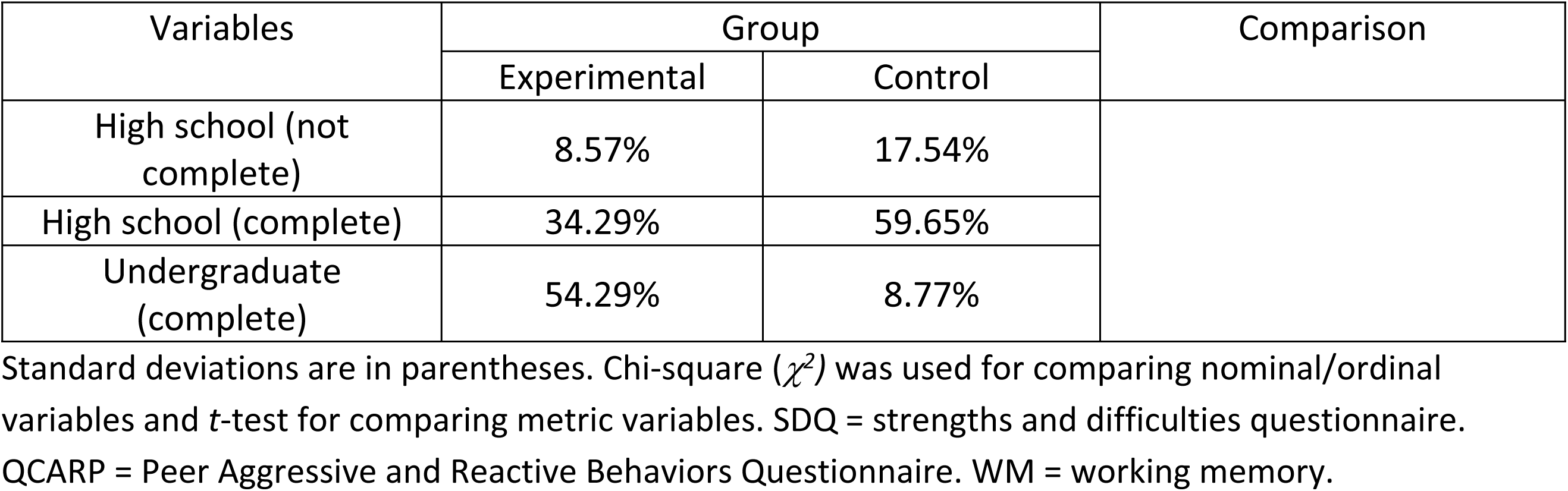
Comparison between groups in terms of baseline characteristics after matching (*n* = 92).

### Statistical analysis

To deal with missing data, imputation was performed. Percent missing for each individual variable varied from 7.1% (times of execution of the Raven’s test; not treated here) from 15.31 (SDQ at the post-test). We replaced missing data using the expectation maximization method with 25 iterations. Little’s MCAR test suggested that missing data was at random (χ^2^(147) = 133.214, *p* = 0.786). We paired the groups using pre-test data (before running the analyses, using data from all children who were paired). By imputing missing data, we did not remove any subjects. Everyone who underwent “treatment” –music classes–, whether absent or present, was part of the clinical group (intention to treat–ITT).

Descriptive statistics–means, standard deviations, and mean differences (between FUP and baseline measures) as well as statistical graphic representations are reported. Inferential analyses are reported by ANOVAS with repeated measures, using group as a fixed factor. Because maternal levels of schooling differed between the groups, it was included as covariate. Interactions between factors, groups, and maternal schooling levels were performed. Data was analyzed using SPSS 23.0 for windows and we consider significant the results with p <0.05. For effect size, *η^2^* is reported, in which: small (*η^2^* < 0.01), medium (*η^2^* between 0.02 and 0.06) and large (*η^2^* > 0.14) effects (Cohen, 2013 [1988]).

## Results

### Behavioral and cognitive results

Table 2 presents the descriptive statistics in the FUP (mean, standard deviation, and means differences between FUP and baseline results). Polled results are also presented (imputation).

**Table 2.** Descriptive statistics of the follow-up measures (pooled means, standard deviation, means differences from baseline)

| Variables | Experimental |  | Control |  |
| --- | --- | --- | --- | --- |
|  | Follow-up | Mean difference (T2-T1) | Follow-up | Mean difference (T2-T1) |
| Age (years) | 5-9 (M = 6.51; sd = 0.61) | 0.475 | 6-8 (M = 6.99; sd = 0.75) | 0.437 |
| SDQ |  |  |  |  |
| Difficulties (total score) | 12.40 (4.85) | -3.08 | 13.66 (6.58) | -1.05 |
| Hyperactivity/Inattention | 5.66 (2.25) | -1.35 | 4.53 (2.63) | -0.71 |
| Peer relationship problems | 2.13 (1.89) | -0.44 | 2.16 (1.76) | -0.33 |
| Conduct problems | 2.27 (1.41) | -0.44 | 2.66 (2.02) | -0.22 |
| Emotional Symptoms | 3.70 (2.15) | -0.85 | 4.30 (2.53) | 0.22 |
| Prosocial behavior (free variation) | 8.06 (1.75) | -0.05 | 8.31 (2.13) | 0.14 |
| QCARP |  |  |  |  |
| Direct aggression | 13.66 (1.66) | 0.43 | 13.45 (2.41) | 1.21 |
| Reactive aggression | 16.12 (2.58) | 0.58 | 15.83 (3.86) | 1.83 |
| Internalizing behavior | 4.98 (2.91) | -0.10 | 4.40 (2.71) | 0.13 |
| Seeking teacher support | 2.12 (2.55) | -0.59 | 2.27 (2.18) | -0.88 |
| WM (reverse span) | 2.69 (0.81) | 0.72 | 2.40 (0.97) | 0.77 |
| Attention |  |  |  |  |
| Sustained Attention | 40.63 (19.30) | 10.09 | 35.84 (20.82) | 11.52 |
| Divided Attention | 33.69 (27.80) | 19.52 | 23.86 (38.00) | 16.84 |
| Alternated Attention | 47.16 (16.07) | 19.82 | 38.77 (15.98) | 14.46 |

Standard deviations are in parentheses. T2 represents follow-up results and T1 baseline results. SDQ = strengths and difficulties questionnaire. QCARP = Peer Aggressive and Reactive Behaviors Questionnaire. WM = working memory.

A repeated-measures ANOVA was conducted to verify the impact of the musical intervention on different variables of the study, considering as factor among subjects the group (control, experimental), schooling as covariate, and observing interactions. For SDQ analyzes, difficulties (*i.e.*, hyperactivity, relationship problems, conduct problems, emotional symptoms, difficulties in general) and prosocial behavior were considered as dependent variables. A reduction in difficulties and an increase in prosocial behavior were the expected outcomes to verify the impact of music education. Baseline and FUP comparisons (ANOVAS) are shown in Table 3.

**Table 3.** Repeated measures comparisons (main effect and interactions) for the behavioral measures (SDQ)^a^.

| Variable (SDQ) | Main effect | Interaction (mother's schooling) | Interaction (group) |
| --- | --- | --- | --- |
| Difficulties (total score) | $F(1,89) = 1.062, p = 0.306, \eta^2 = 0.012$ | $F(1,89) = 0.047, p = 0.829, \eta^2 = 0.001$ | $F(1,89) = 3.179, p = 0.078, \eta^2 = 0.034$ |
| Hyperactivity/Inattention | $F(1,89) = 0.461, p = 0.499, \eta^2 = 0.005$ | $F(1,89) = 0.062, p = 0.084, \eta^2 = 0.001$ | $F(1,89) = 1.185, p = 0.279, \eta^2 = 0.013$ |
| Peer relationship problems | **$F(1,89) = 6.615, p = 0.012, \eta^2 = 0.069$** | **$F(1,89) = 4.431, p = 0.0338, \eta^2 = 0.047$** | $F(1,89) = 1.280, p = 0.261, \eta^2 = 0.014$ |
| Conduct problems | $F(1,89) = 0.048, p = 0.828, \eta^2 = 0.001$ | $F(1,89) = 0.376, p = 0.542, \eta^2 = 0.004$ | $F(1,89) = 0.108, p = 0.744, \eta^2 = 0.001$ |
| Emotional Symptoms | $F(1,89) = 0.014, p = 0.907, \eta^2 = 0.000$ | $F(1,89) = 0.040, p = 0.841, \eta^2 = 0.000$ | **$F(1,89) = 4.562, p = 0.035, \eta^2 = 0.049$** |
| Prosocial behavior (free variation) | $F(1,89) = 0.164, p = 0.687, \eta^2 = 0.002$ | $F(1,89) = 0.230, p = 0.632, \eta^2 = 0.003$ | $F(1,89) = 0.568, p = 0.442, \eta^2 = 0.007$ |

<sup>a</sup>SDQ = strengths and difficulties questionnaire.

As seen in Table 3, the main effect was significant for peer relationship problems, indicating that both groups reduced these symptoms between baseline and FUP testing (see difference means in Table 2). There was a significant interaction between peer relationship problems and maternal schooling levels; the decrease in relationship problems was greater depending on the schooling level of the mother, irrespective of group. Post hoc comparisons showed that children whose mothers had medium to high years of education tended to start the study with higher levels of peer relationship problems, compared to children whose mothers had lower levels of schooling. Yet, at FUP their scores were equivalent to those of children with less educated mothers (see S3 Table). The decrease in scores for relationship problems was greater for children with mothers who did not complete high school, although the lowest score for relationship problems (mean 1.19) came from children with mothers who had attended elementary school. In terms of group interactions, significance was found only for emotional symptoms. While mean scores for emotional symptoms decreased for the experimental group at FUP, they had a slight increase for controls. As seen in Table 2 (difference means from baseline to FUP) and Fig 2, emotional symptoms fell among the experimental group and tended to increase slightly in the control group.

**Fig 2.**
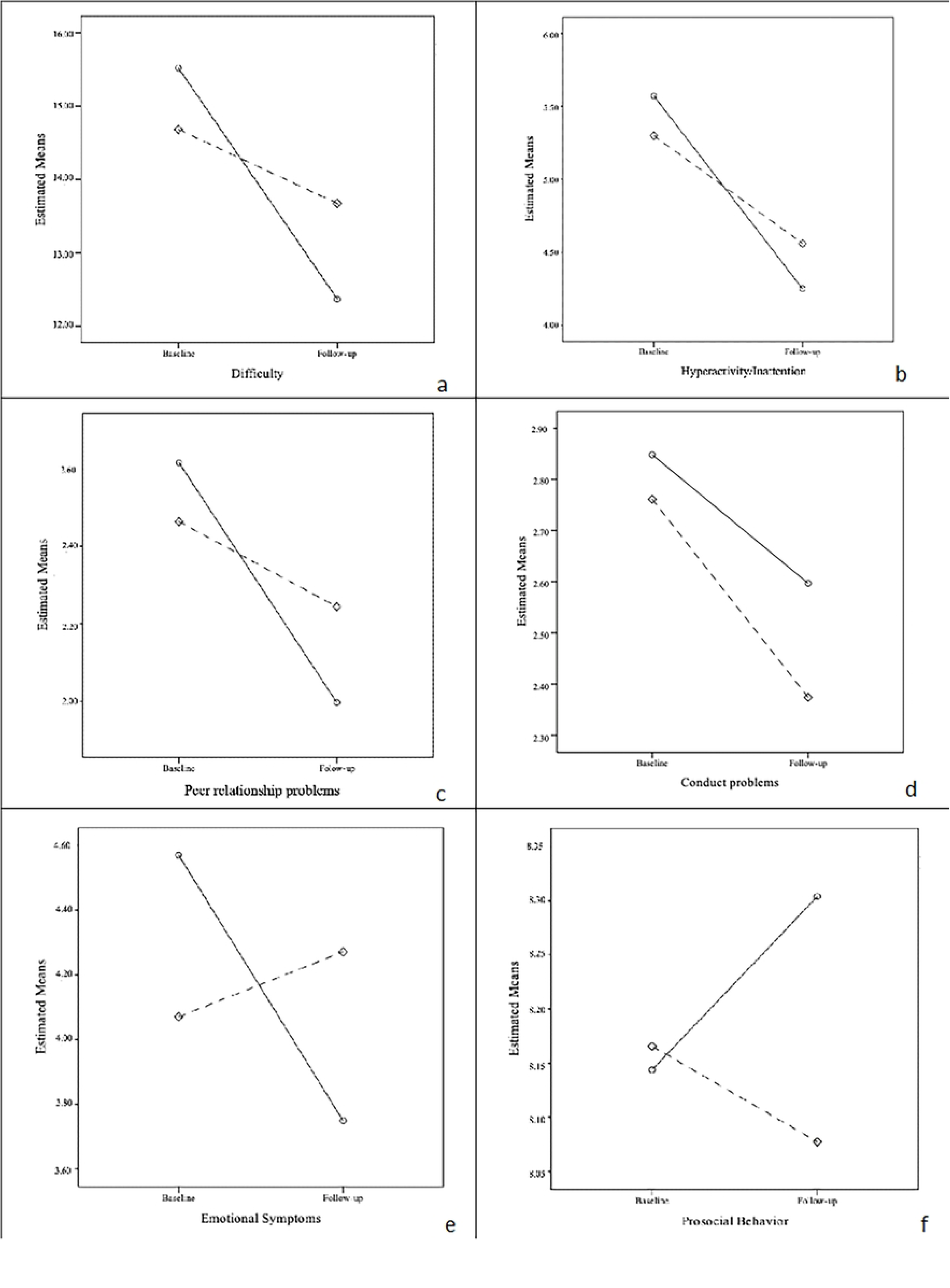
Comparative charts of SDQ symptoms and behaviors in the baseline and follow up. Solid lines represent the experimental group and dashed lines the control group. Scales are not standardized and represents estimated means generated for the program by default.

Comparisons between groups for direct aggressive behavior and reaction to aggression assessed by the QCARP are shown in Table 4. No significant effects were found for any of the aggression measures.

**Table 4.** Repeated measures comparisons (main effect and interactions) for the behavioral measures (aggression – QCARP)

| Variable (QCARP) | Main effect | Interaction (mother's schooling) | Interaction (group) |
| --- | --- | --- | --- |
| Direct aggression | $F(1,89) = 0.229, p = 0.633, \eta^2 = 0.003$ | $F(1,89) = 0.014, p = 0.905, \eta^2 = 0.000$ | $F(1,89) = 1.529, p = 0.220, \eta^2 = 0.017$ |
| Reactive aggression | $F(1,89) = 1.735, p = 0.633, \eta^2 = 0.003$ | $F(1,89) = 0.014, p = 0.905, \eta^2 = 0.000$ | $F(1,89) = 1.529, p = 0.220, \eta^2 = 0.017$ |
| Internalizing behavior | $F(1,89) = 0.092, p = 0.762, \eta^2 = 0.001$ | $F(1,89) = 0.102, p = 0.750, \eta^2 = 0.001$ | $F(1,89) = 0.173, p = 0.678, \eta^2 = 0.002$ |
| Seeking teacher support | $F(1,89) = 1.061, p = 0.306, \eta^2 = 0.012$ | $F(1,89) = 0.358, p = 0.551, \eta^2 = 0.004$ | $F(1,89) = 0.021, p = 0.884, \eta^2 = 0.000$ |

QCARP = Peer Aggressive and Reactive Behaviors Questionnaire.

Fig 3 depicts the baseline and FUP results for these measures.

**Fig 3.**
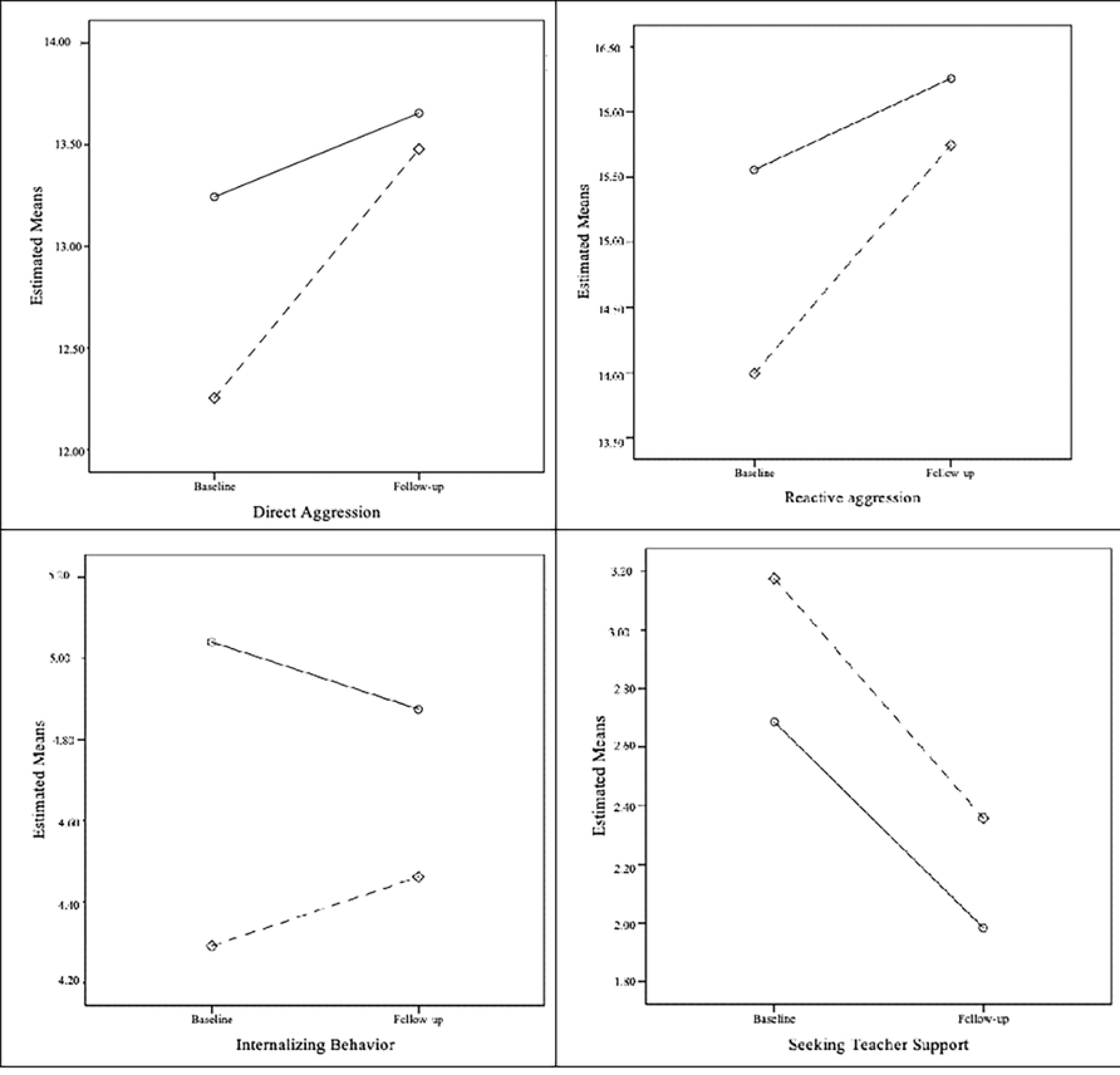
Comparative charts of aggressive behavior and reaction to aggression evaluated by the QCARP in the baseline and follow-up. Solid lines represent the experimental group and dashed lines the control group. Scales are not standardized and represents estimated means generated for the program by default.

The results for verbal working memory assessed by digits span (reverse order) and the attentional measures are shown in Table 5 (and their respective descriptive statistics in Table 2).

**Table 5.** Repeated measures comparisons (main effect and interactions) for the cognitive measures.

| Variable | Main effect | Interaction (mother's schooling) | Interaction (group) |
| --- | --- | --- | --- |
| WM<sup>b</sup> (reverse span) | $F(1,89) = 2.443, p = 0.122, \eta^2 = 0.027$ | $F(1,89) = 0.072, p = 0.789, \eta^2 = 0.001$ | $F(1,89) = 0.006, p = 0.936, \eta^2 = 0.000$ |
| Sustained Attention | $F(1,89) = 2.985, p = 0.088, \eta^2 = 0.032$ | $F(1,89) = 0.885, p = 0.349, \eta^2 = 0.010$ | $F(1,89) = 0.020, p = 0.888, \eta^2 = 0.000$ |
| Divided Attention | **$F(1,89) = 14.221, p < 0.001, \eta^2 = 0.032$** | **$F(1,89) = 5.476, p = 0.006, \eta^2 = 0.083$** | $F(1,89) = 2.000, p = 0.161, \eta^2 = 0.022$ |
| Alternated Attention | **$F(1,89) = 9.853, p = 0.002, \eta^2 = 0.100$** | $F(1,89) = 2.605, p = 0.110, \eta^2 = 0.028$ | $F(1,89) = 2.747, p = 0.101, \eta^2 = 0.030$ |

<sup>b</sup>WM = working memory.

As shown in Table 5, there were significant main effects for divided and alternated attention. For both groups, attention scores increased from baseline to FUP. There was a significant interaction between maternal schooling levels and divided attention. Post hoc analyses showed that children with mothers who had low levels of education benefited more from the development of divided attention, with gains varying around 0.70 to 0.95 standard deviations (S4 Table). No other significant interactions emerged for any of the cognitive variables (Fig 4).

**Fig 4.**
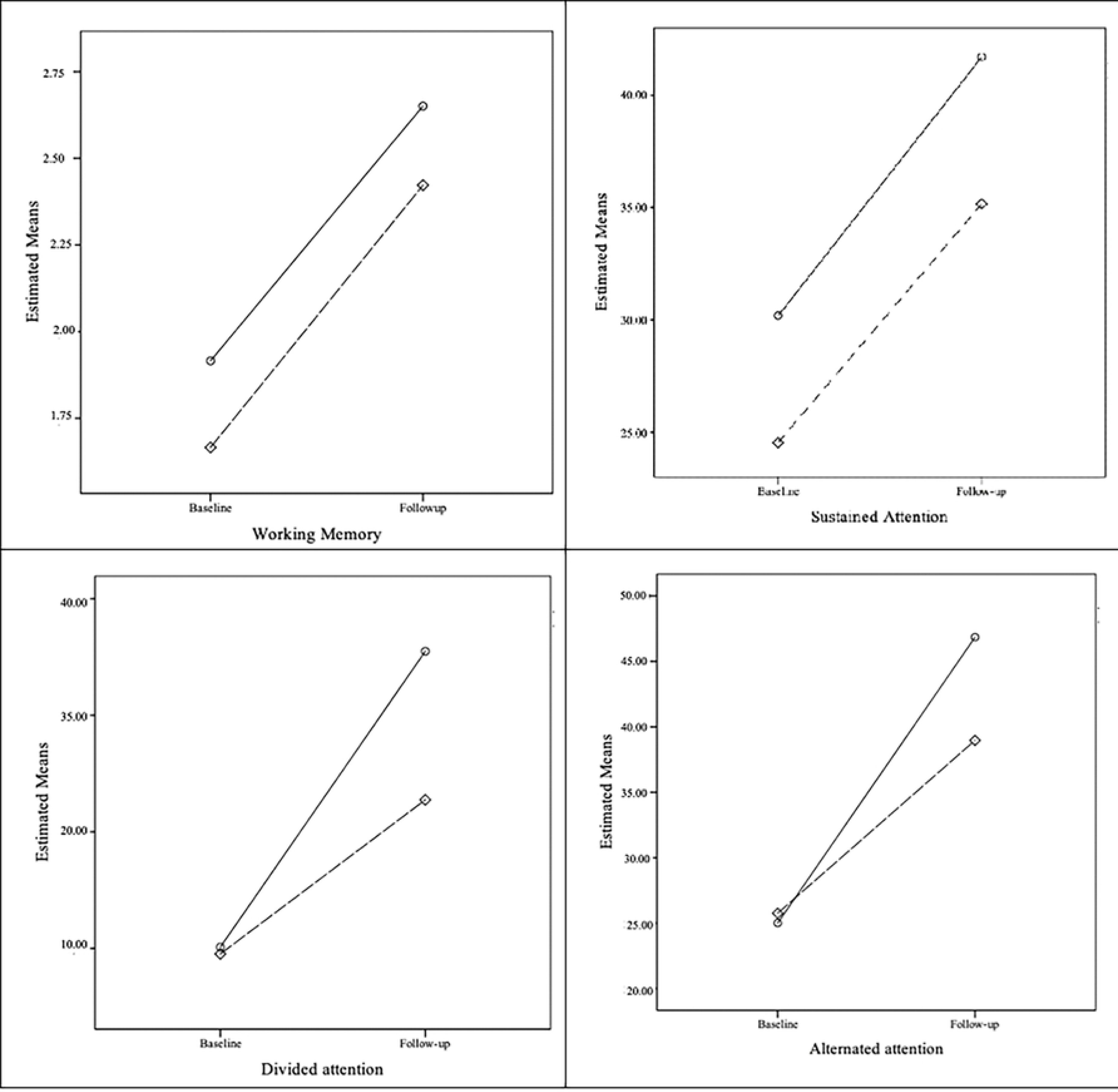
Comparative charts for the cognitive measures (working memory and attention) in the baseline and follow-up. Solid represent the experimental group and dashed lines the control group. Scales are not standardized and represents estimated means generated for the program by default.

## Discussion

The aim of this quasi-experimental study was to examine the impact of a music education program on aspects of attention—shifting, divided, and sustained—, socioemotional abilities, and reactive and aggressive behaviors in children aged 5-9 from underserved communities of São Paulo. Using standardized tests, we compared results from a group of children who participated in a community-based music program to matched controls at two time points: before the intervention and seven months later. Because maternal education is known to have an effect on children’s cognitive and socioemotional development (62–64), we also analyzed maternal education data reported by the children’s parents through the ABEP socioeconomic status questionnaire (52). To our knowledge, this is likely the first quantitative study carried out on the impact of sociomusical programs in Brazil.

A main finding that emerged from our study was a significant effect of the intervention on the music group in the FUP. Our data revealed a more pronounced decrease in emotional symptoms in the music group as opposed to controls. This finding is consistent with earlier theoretical (65) and empirical studies on the role of music education in children’s mood and emotional regulation (66, 67). We also found a developmental effect for emotional symptoms, with children in both groups showing improved scores from baseline to FUP. Emotional symptoms including depression, anxiety, perceived peer rejection, victimization, and feelings of loneliness, have been associated with motor competence and to one’s ability to move with rhythm (67–69). It has been established that motor coordination disorders are risk factors for mental health problems, such as eating disorders (68, 70, 71). Considering that rhythmic training is intrinsic to music education in the Guri’s program, we suspect that it may have been a protective factor for children’s emotional symptoms.

In this study, maternal schooling was found to interact with relationship problems and emotional symptoms in different ways. Children of more educated mothers had more relationship problems at the start of the study in both groups, but these balanced out after 7 months. Emotional symptoms decreased more markedly in children whose mothers had less education. Why would maternal education impact these two socioemotional areas in such distinct ways? Previous studies (63) have found higher maternal education levels (*i.e.*, at least some college education) to be associated with fewer behavioral problems in children, although earlier comparisons of different levels of maternal education (*i.e.*, without some college education) revealed similar results from those of our study. Maternal education has been linked to numerous practices that grant children cognitive advantages, such as the use of more complex language (63). As socioemotional symptoms were assessed through parental reports, it is possible that children with more educated mothers were also more prone to verbalize the impact of formal learning environments in public schools as opposed to the home environment in the aftermath of the pandemic. Our study took place immediately after the social restrictions imposed by the coronavirus pandemic began to be lifted. In Brazil, there also were marked differences in access to education between families during the COVID-19 quarantine. This is, of course, speculative. By contrast, the finding of a significant interaction between maternal schooling and children’s emotional symptoms is consistent with earlier studies that found a close relationship between maternal education and children’s mental health (62). Maternal education has been directly associated with low SES and social status, another factor to have an impact on children’s mental health and socioemotional wellbeing (72), particularly in early childhood (62).

In terms of cognitive outcomes, divided attention had a developmental effect, increasing equally between groups, from baseline to FUP. We also found divided attention to increase more in children whose mothers had lower levels of education. A possible explanation for this finding is that in formal educational settings, children from our study were more able to learn and retain new information, keeping up with other children’s skills. During the pandemic, many children spent long periods of time in front of screens (*e.g.*, TV, tablets, games, and cell phones), which is known to reduce their attention span (73). Furthermore, children from low-SES families in Brazil were often left at home unsupervised (74). Alternated attention, in turn, was higher for the music group, although no causal relationship was observed. Contrary to earlier studies, we did not find effects of music instruction on children’s attention, possibly due to our small sample size, the short duration of the intervention (*i.e.*, 7 months) and music classes (*i.e.*, only one hour per week), and the unprecedent pandemic variable.

## Conclusion & Limitations

Parents of the children selected for this study reported high difficulties in the SDQ, which suggests a risk of exposure to mental health vulnerabilities. The surveys were carried out in peripheral regions of São Paulo, Brazil, where services such as public security, health, education, and transportation are scarce and oftentimes precarious where they exist. The literature indicates such elements as predictors of emotional symptoms, health problems related to emotional stress, and behavioral problems in children (75).

Given the harmful effect of scarcity on mental health of underserved communities, we believe it is crucial that public policies pay attention to the potential benefits of music education on socioemotional symptoms. It is essential that the State, public policy administrators, as well as the third sector represented by social organizations and non-governmental organizations (NGOs), pay attention to the responsibility of expanding the scope of their activities to these vulnerable populations. Access to music education can potentially serve as a protective factor for children exposed to violence and cognitive vulnerabilities.

As limitations of this study, the absence of statistically significant interaction between the groups in some cases may be due to the low sample size and short intervention time. Also, a question remains unanswered is: Would different curricular approaches to music education yield different results? Previous studies suggest that curated curricula may yield more substantial findings in terms of both EFs (76) and socioemotional skills (77). Additionally, the length of programs is important; the literature points to the potentiation of long-term exposure to music practice (more than two years) on the impact of behavioral measures and brain structures (3, 8, 78–80). Our study collected data over a school year, which, in practice, had an interval between the baseline and FUP of only about seven months, and included an hour of music class per week, and in the aftermath of a global pandemic. We hope that the tendency observed here can contribute to future research. More studies are obviously needed.

## Data Availability

We have a restriction imposed by the Consent Form signed by both researchers and parents that affirms: “The researchers are committed to using the data collected only for this research.” In case it is strictly necessary to check the data, they are available from the Ethical Committee Plataforma Brasil n. 15049919.8.0000.5420 at https://plataformabrasil.saude.gov.br

## Acknowledgements

We thank all the families and their children who volunteered in this research, as well as the assistants who collected the data, the Projeto Guri (OS Santa Marcelina), Secretaria Municipal de Educação de São Paulo, and Secretaria da Cultura e Economia Criativa do Estado de São Paulo for granting permission for the study to take place.

## Supporting information

**S1 Table. Comparison of groups in terms of baseline characteristics.** *Note*. Standard deviation in parentheses. Chi-square *(χ^2^)* for comparing nominal/ordinal variables and t-test for comparing metric variables. SDQ = strengths and difficulties questionnaire. Qcarp = Peer Aggressive and Reactive Behaviors Questionnaire. PA = active aggression; RA

**S2 Table. Matching according to age, levels of reaction to aggression and performance in the Raven test.** *Note*. RA = reactive aggression in Q-carp questionnaire. Intelligence in Raven test was classified as follows: 1 = superior; 2 = Medium superior; 3 = Medium; 4 = Medium Inferior; 5 = Inferior (not present in the sample; removed from inclusion criteria). Mother’s schooling was classified as follows: 1= illiterate; 2 = Elementary school (incomplete/complete); 3 = High school (not complete); 4 = High school (complete); 5 = Undergraduate (complete).

S3 Table. Descriptive statistics for the peer relationship problems considering mother’s level of education. *d* = Cohen’s d (0.30 = low; 0.5 moderate; 0.80 high).

S4 Table. Descriptive statistics for the peer relationship problems considering mother’s level of education. *d* = Cohen’s d (0.30 = low; 0.5 moderate; 0.80 high).

## Footnotes

1 The data discussed here were first presented as a thesis of senior professor exam in 2023 by the corresponding author. It remains unpublished, and the results were reviewed in this article.

2 The children also underwent MRI scans at baseline and follow-up. These data are not included in this article. For adherence, the children then received a gift, a snack, a certificate of contribution to scientific research and their families received a refund for transportation.

3 The original schedule proposed in the trial study protocol predicted that the intervention would take place during the 2020 academic year, preceded by a pilot study in 2019. Due to the Covid-19 pandemic, the intervention had to be interrupted during the initial data collection, in March 2020, and the study was resumed in 2022 with new participants.

4 In the trial study protocol, we intended to test children aged between 6 and 7, but as it was difficult to achieve the minimum sample size desired in this group, the range was extended.

5 This study started at 2020, but when the intervention was about to begin, the peak of the coronavirus pandemic come. By that time, although the baseline assessment had already finished, the children of the experimental group were taking remote music classes and the research had to be cancelled. Therefore, this sample is different from that in their pre and post-test assessments.

